# Adolescent satisfaction with public health services and contraceptive use in Nepal - A sequential explanatory mixed methods study

**DOI:** 10.64898/2026.05.04.26352425

**Authors:** Santa Kumar Dangol, Megh Raj Dangal, Sujan Babu Marahatta, Adweeti Nepal

**Affiliations:** Kathmandu University, School of Arts, Department of Development Studies, Balkumari, Lalitpur, Nepal; Medical Education Commission, Sanothimi, Bhaktapur, Nepal; Center for International HealthLMU, Munich, Germany

**Keywords:** Adolescents, contraceptive use, perceived satisfaction, adolescent-friendly health services and health-service delivery

## Abstract

**Background:** Limited access to and use of contraceptive services among adolescents remain a major public health concern in Nepal, influenced by their experiences and satisfaction with health services. Understanding the factors that influence adolescents’ satisfaction with health services is essential for improving access to and utilization of contraceptive services. This study explores determinants of adolescents’ satisfaction with health services and how these factors influence contraceptive service use in Nepal.

**Method:** An explanatory sequential mixed-methods design was employed in 154 health facilities across randomly selected 28 local levels in six districts (Surkhet, Banke, Pyuthan, Nuwakot, Parsa, and Siraha) of Nepal. Quantitative data were collected through client-exit interviews with154 adolescents on their health facility visit day, followed by qualitative interviews. Total 12 focus group discussions were conducted with adolescent girls and boys. Quantitative data were analyzed using SPSS version 26, while qualitative data were transcribed, systematically coded, and analyzed using deductive thematic approach.

**Finding:** In quantitative results, it is found that overall, 82.5% of adolescents reported satisfaction with health services on the day of visit. The key health system factors were significantly associated with satisfaction, including confidentiality (AOR: 3.50; 95% CI: 1.19–10.37) and ease of obtaining appointments (AOR: 6.28; 95% CI: 2.18–18.08). No significant association were observed between satisfaction and adolescent’s socio-demographic characteristics. Despite the high-level satisfaction reported in quantitative interviews, qualitative findings revealed contrasting experiences. Adolescents reported issues such as providers judgmental attitude, inadequate confidentiality and privacy, discriminatory behavior, and limited participation in decision-making processes, influencing their service seeking behavior from public health facilities.

**Conclusion:** This study highlights the central role of health system factors in shaping adolescents’ satisfaction with and use of contraceptive services. Strengthening these dimensions is essential to improve contraceptive uptake among adolescents in Nepal.

## Introduction

Adolescence (10–19 years) is a crucial developmental phase of human life marking the transition from childhood to adulthood [1]. Globally, 1.3 billion adolescents account for 16% of the population, representing the largest generation in history [2]. In this critical stage, adolescents go through several physiological, mental, and social changes, including those related to sexual and reproductive health and development [1]. A significant concern worldwide within this demographic is early pregnancy among adolescent girls. Approximately 13% of girls give birth before reaching the age of 18, which subjects them to considerable health, social, and economic risks [3,4]. Early childbearing adversely affects their access to education, constrains future economic opportunities, perpetuates gender inequalities, and contributes to their long-term vulnerability [5,6].

In Nepal, adolescents account for 20.1% of the population with girls comprising half of this group [7]. Yet, early pregnancy remains widespread, by age of 19, around 17% of girls have already begun childbearing [8]. This not only expose young girls to significant health risks but also contributes to 10% of maternal deaths in the country, underscoring the urgency of addressing their reproductive health needs [9]. Although contraception is a proven strategy to prevent unintended pregnancies, access to and use among adolescents remained low, only 14% of married adolescents use modern methods, while nearly one-third have an unmet need [5]. As a result, the adolescent fertility rate remains high at 71 births per 1,000 girls, with minimal progress over the past two decades [8,10].

Satisfaction with health services is a critical yet often overlooked determinant of adolescents’ use of contraceptive services. Their decisions to seek, initiate, and continue using contraception are shaped by their experiences within the health system [11]. Evidence indicates that adolescents are more likely to utilize services when health workers demonstrate respectful, non-judgmental attitudes and ensure privacy and confidentiality [10–12]. In addition, provider competence, communication skills, and counseling quality influence adolescents’ access to and choice of contraceptive methods [13]. In response to persistent quality gaps, the Government of Nepal has implemented the adolescent friendly health services through health facilities focusing on responsive and accessible sexual and reproductive health (SRH) services, that emphasizes confidentiality, privacy, non-discriminatory care, access to information, and meaningful adolescent participation in health planning and decision-making process [14,15]. To further improve access, government has established a comprehensive legal framework that safeguards the sexual and reproductive health and rights (SRHR) of all women including adolescent girls and guarantees access to information and services related to contraception [16].

While previous studies in Nepal have explored barriers to access and use of adolescent sexual and reproductive health (ASRH) and contraceptive services among adolescent and young women, they have largely focused on structural and socio-cultural constraints in a broader sense [12,17,18]. However, limited evidence exists on how adolescents’ satisfaction with the health services they receive influences their health-seeking decisions, particularly their willingness to use contraceptive services. Specifically, health-system factors that shape satisfaction and subsequently affect adolescents’ contraceptive behaviors remain insufficiently explored in the Nepali context. This gap underscores the need for a more nuanced understanding of how service experiences, beyond access alone, guide adolescents’ contraceptive decision-making.

To address this knowledge gap, this study aims to explore the determinants of adolescents’ satisfaction with public health services and examines how these determinants relate to their contraceptive use in the districts with high and rising adolescent pregnancy rates in Nepal [19]. Using a sequential mixed-methods approach, the study integrates quantitative and qualitative data to generate a comprehensive understanding of adolescents’ service experiences, offering insights that can inform policies and interventions aimed at strengthening adolescent-friendly health services.

## Methodology

### Study design and setting

A sequential explanatory mixed-methods design [20] was employed to examine adolescents’ satisfaction with health services and their relationship with contraceptive use. This approach involved an initial quantitative phase followed by a qualitative phase, where qualitative findings were used to explain and contextualize the quantitative results.

### Conceptual framework

The conceptual framework used in this study is guided by the Andersen Behavioral Model of Health Service Utilization, which was originally developed in the 1960 [21]. The model proposes that individuals’ use of health services is shaped by three key components: their predisposition to seek care, the enabling or limiting resources that facilitate or restrict access, and their perceived or actual need for healthcare. In this study, adolescents’ use of health services particularly contraceptive services is understood through the interaction of predisposing socio-demographic factors, enabling health-system factors, and their service experiences. Socio-demographic characteristics such as age, gender, education, marital status, religion, caste/ethnicity, and place of residence serve as independent variables, highlighting variations in access and satisfaction. Health-system elements include the availability of information, education and communication (IEC) materials, confidentiality, non-discriminatory treatment, ease of obtaining appointments, and adolescent participation function as intermediate factors that shape adolescents’ experiences of care. Satisfaction with health services reflects how adolescents perceive these interactions and ultimately shapes their likelihood of using contraceptive services (Table 1 for operational definition).

**Table 1:** Study variables and operational definitions.

| Category | Variables | Operational Definitions |
| --- | --- | --- |
| **Service Utilization** | Contraceptive service use | Use of counselling service and contraceptive services |
| **Outcome Variable** | Satisfaction with service of the visited health facility | Reported satisfaction by adolescents with the health services received from the health facility on the day of their visit. Labeled with satisfaction and unsatisfaction. |
| **Intermediate Variables** | Availability of Information Education and Communication (IEC) materials | Whether IEC materials were available at the health facility at the time of service or visit. |
|  | Non-discriminatory service | Adolescents' perception of receiving fair, respectful, and non-biased treatment from health workers. |
|  | Confidentiality of information | Whether adolescents perceived that their personal information is kept private by the health facility. |
|  | Appointment with health workers | Ease to obtain an appointment or being able to meet with a health worker when needed. |
|  | Adolescent participation in health system | Adolescents' awareness of, or involvement in, decision-making and planning processes at health facility level and in health facility operation and management committee (HFOMC). |
| **Independent Variables** | Place of residence | Local level of residence of respondents (Municipality or Rural Municipality). |
|  | Type of local level | Municipality and Rural municipality |
|  | District | • Surkhet (Karnali Province)<br>• Banke (Lumbini Province)<br>• Pyuthan (Lumbini Province)<br>• Nuwakot (Bagmati Province)<br>• Parsa (Madhesh Province)<br>• Siraha (Madhesh Province) |
|  | Age | Age groups 16, 17, 18, and 19 years. |
|  | Gender | Male or Female. |
|  | Marital status | Married or Unmarried at the time-of-service use and interview. |
|  | Education level | • Basic (Grade 1–8),<br>• Secondary Education Examination (up to grade 10)<br>• Higher Secondary level (Grade 11–12). |
|  | Type of health facility | Basic hospital, Primary Health Care Center (PHCC), Health Post, Basic Health Care Center. |
|  | Caste/Ethnicity | 1. Brahmin/Chhetri: This group includes Hill Brahmin/Chhetri and Terai Brahmin/Chhetri.<br>2. Dalit: This group includes Hill Dalit and Terai Dalit.<br>3. Janjati: This group includes Newar and both Hill and Terai Janajati.<br>4. Others: This category includes Muslims, Terai other castes, and other caste groups. |

### Study setting and sampling

The study was conducted in six districts Surkhet, Banke, Pyuthan, Nuwakot, Parsa, and Siraha of Nepal, selected purposively, considering high and increasing adolescent fertility rate trends ensuring geographic (hill and *Terai*^1^ regions), socio-cultural and ethnic diversity [19]. The level of adolescent satisfaction with health services was unknown; therefore, a prevalence of 50% was assumed for the sample size calculation to ensure maximum variability. After adjusting for the design effect associated with multistage sampling and inflating for a 7% non-response rate, the final calculated sample size of the study was 154 as below[22]. Data were collected by interviewing one adolescent client from each basic healthcare facility on the day of the visit.

From a total of 69 local levels across these six districts, 28 local levels were selected using a computer-generated random number table in Microsoft Excel [23] to get sufficient number of adolescent assuming 5-6 health facilities providing health service to adolescent on the day of visit.

### Sample size calculation

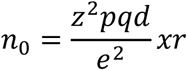

Where,

q= 1-p
z=1.96 (at 95% level of confidence)
p = 0.5 (Maximum variability taken; as standard for unknown prevalence)
d=1.5 (design effect for multistage sampling)
e = 0.20 (relative margin of error)
r= 7% (nonresponse rate)

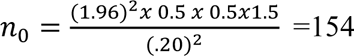

A margin of error of 20% was considered, given the explanatory sequential mixed-methods design, which prioritized feasibility over strict statistical precision. In this context, qualitative findings were used to complement quantitative results, allowing flexibility in precision requirements appropriate for a small-scale study [20,24].

**Figure 1:**
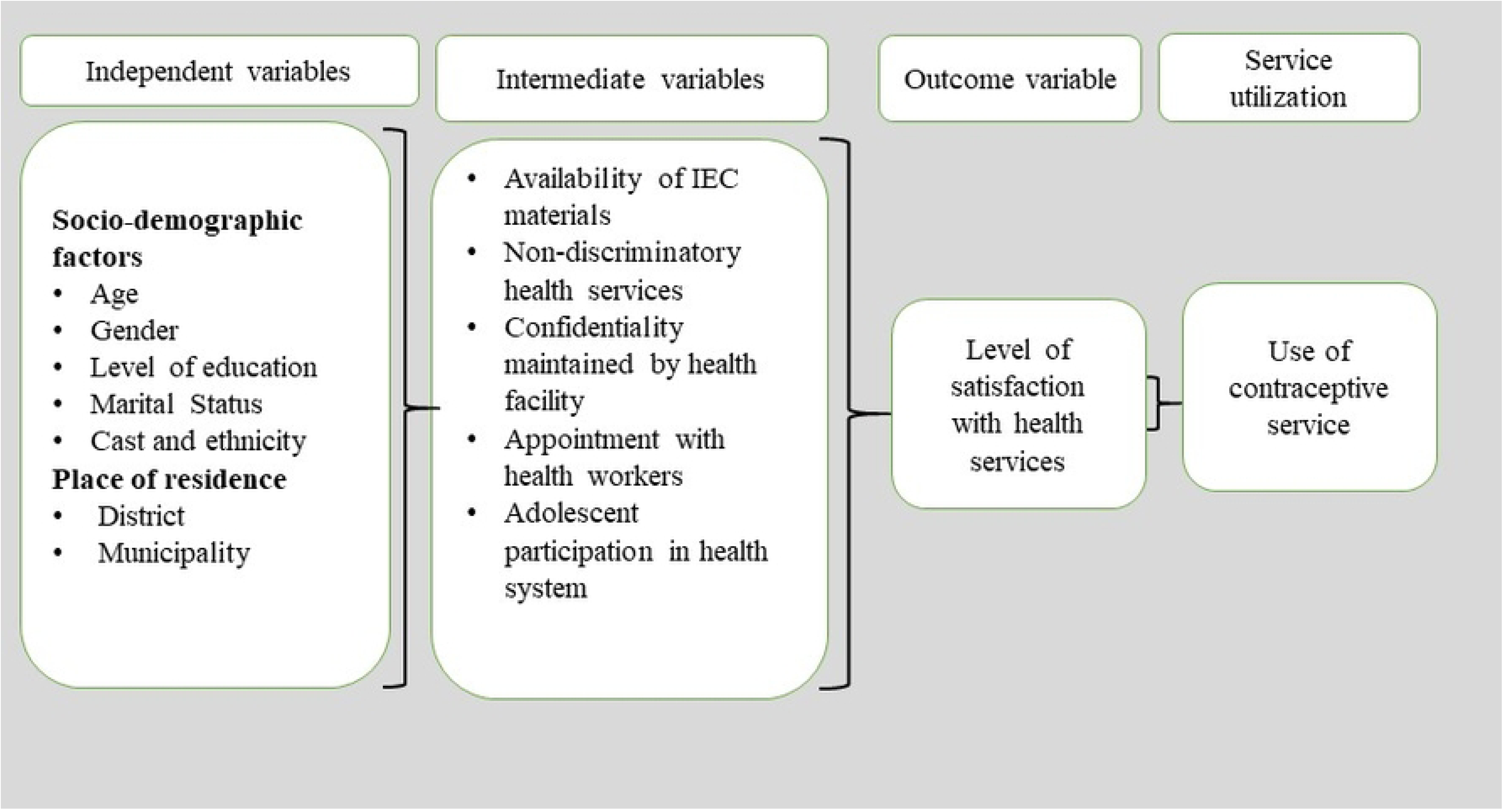
Conceptual framework adapted from Andersen Behavioural Model [21]

### Data Collection

#### Quantitative data collection

In the first phase, the study collected quantitative data from April 2025 to June 2025 through structured client-exit interviews with adolescents (Figure 2). Data collection was conducted at each facility during outpatient service hours (10:00 am to 2:00 pm, Sunday to Friday). Adolescents who visited the health facility on the day of data collection and provided consent to participate were included in the study.

**Figure 2:**
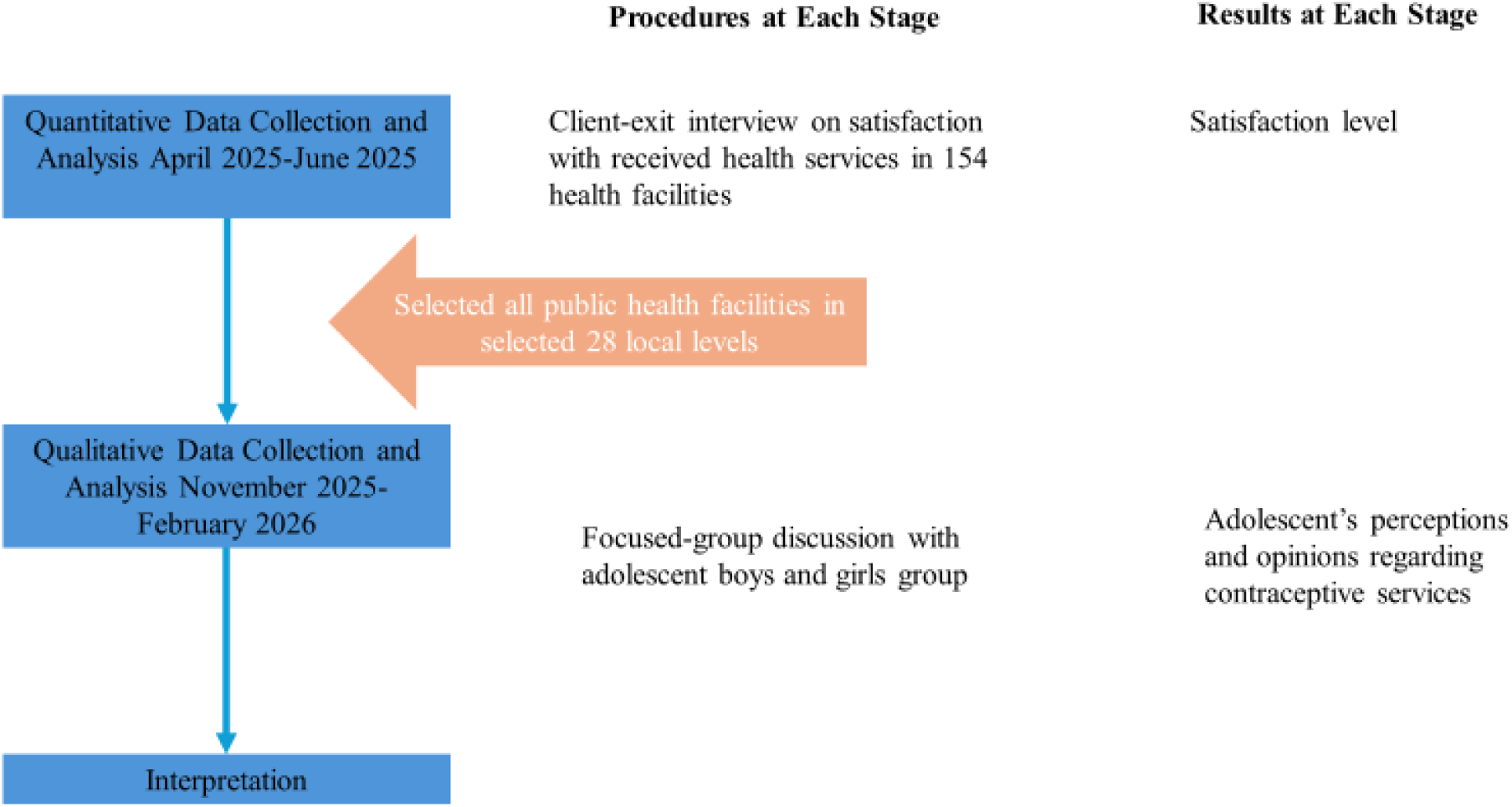
Explanatory sequential design of study[20].

Although adolescents aged 15–19 years are generally considered the target group for SRH services, only those aged 16–19 years were identified and included during data collection based on self-reported age at the time of interview. Adolescents who were severely ill or unwilling to participate were excluded. A total of 154 adolescents who sought any type of health service during the visit were available and included as respondents for client-exit interviews.

The exit interview questionnaire was developed based on the quality measurement standards outlined in the Adolescent Friendly Health Service guideline (2022) developed by Family Welfare Division [15]. The tool was developed using a Likert scale to measure the level of satisfaction. The tool pre-tested prior to data collection, and necessary revisions were made. Enumerators were recruited and trained on study procedures, ethical considerations, and standardization of interview techniques to ensure data quality and consistency before field deployment.

#### Qualitative data collection

In the qualitative data collection phase, 12 focus group discussions (FGDs) were conducted separately with adolescent girls (n = 60) and boys (n = 59) aged 15–19 years (Table 2) in period between November 2025-February 2026 (Figure 2). Participants were purposively selected from the study sites ensuring diversity in age, sex, and socio-cultural background. Each FGD consisted of approximately 8–12 participants and lasted between 60–90 minutes. Discussions were conducted by first author (SKD) himself for male group, while female enumerator and co-author (AN) conducted FGD with female groups supported by note-takers. The FGD guide was developed based on key findings from the quantitative phase and explored adolescents’ perceptions, experiences, and barriers related to health service utilization, particularly contraceptive services. Moreover, local enumerators were also hired for Parsa and Siraha district to facilitate the FGDs with adolescents in local language.

**Table 2:** Details of FGD participants.

| Category | Surkhet | Banke | Pyuthan | Nuwakot | Parsa | Siraha | Total |
| --- | --- | --- | --- | --- | --- | --- | --- |
| 6 FGDs with Male Adolescents | 8 | 8 | 8 | 11 | 12 | 12 | 59 |
| 6 FGDs with Female Adolescents | 10 | 8 | 9 | 9 | 12 | 12 | 60 |
| Total | 18 | 16 | 17 | 21 | 24 | 24 | 119 |

### Data Analysis

#### Quantitative data analysis

Quantitative data were entered, cleaned, and analyzed using SPSS version 26. Descriptive statistics were used to summarize participant characteristics and satisfaction levels. For analysis; data collected using the Likert scale categorized the responses of “satisfied” and “very satisfied” as “satisfied,” while all other three types responses were classified as “unsatisfied.”. Binary and multivariate logistic regression analyses were conducted to examine associations between adolescents’ satisfaction with health services (outcome variable) and socio-demographic and health system factors (independent variables). Crude/Adjusted odds ratios (COR and AOR) with 95% confidence intervals (CI) were calculated, and statistical significance was set at p ≤ 0.05.

#### Qualitative data analysis

All FGD notes and audio recordings were transcribed verbatim and translated from Nepali into English by the first author (SKD), who is fluent in both English and Nepali. To ensure translation accuracy and consistency, the English transcripts were independently back translated by another co-author (AN). The back-translated versions were compared with the original Nepali transcripts, and any discrepancies in meaning were discussed and resolved within the research team to ensure conceptual equivalence. The transcribed data were anonymized and securely stored on a password-protected computer accessible only to the research team.

Trustworthiness of the data was ensured through multiple strategies. Methodological triangulation was achieved by conducting FGDs with diverse groups of participants, including male and female adolescents from different settings. In addition, an independent qualitative researcher reviewed the coding process and preliminary analysis to enhance confirmability and reduce potential researcher bias.

Qualitative data were coded and managed manually and analyzed using a deductive thematic approach[25]. Preliminary analysis was carried out concurrently with data collection, allowing emerging findings to inform subsequent data collection and ensure comprehensive exploration of the research questions. Final and systematic analysis was conducted after the completion of data collection.

The analysis followed the six-step framework of Braun and Clarke[26], including transcription and familiarization through repeated reading of transcripts, generation of initial codes, searching for patterns and meanings in the data, reviewing and refining codes, developing and defining themes, and finalizing the analytic framework. Initial coding was done by first author SKD and subsequently discussed with the other authors to reach consensus. A codebook with definitions was developed and iteratively refined as new codes emerged during analysis.

Through a deeper interpretive process, codes were further examined to identify underlying meanings beyond the surface-level content of participants’ narratives. The codes were compared within and across participant groups to identify patterns and variations, which were then synthesized into broader analytical themes aligned with the study objectives. The analysis was guided by the study’s conceptual framework, and illustrative quotations were used to support and substantiate the key findings.

### Integration of findings

Findings from both quantitative and qualitative phases were integrated at the interpretation stage. Qualitative results were used to explain, expand, and contextualize quantitative findings, providing a comprehensive understanding of adolescents’ satisfaction with health services and its influence on contraceptive use (Table 6).

### Ethical consideration

Ethical approval for the study was obtained from the Nepal Health Research Council (Ref. No. 164-2025). Additional permission was secured from the relevant provincial and local authorities, as well as the participating health facilities, after providing them with a brief overview of the study objectives. Written informed consent was obtained from adolescents aged 18 and 19 years, while written assent was obtained from the parents/guardians of adolescents aged 16–17 years prior to the interviews. The purpose of the study, data collection procedures, potential benefits, confidentiality measures, and the voluntary nature of participation including the right to withdraw at any time were clearly explained during the consent and assent process. Participants were assured that the information they provided would remain confidential.

## Results

### Quantitative Findings

A total of 154 adolescents; one from each health facility participated in the client-exit interviews across the six study districts. Most respondents were from Health Posts (70.1%), followed by Basic Health Care Centers (22.1%), PHCC (10; 6.5%), and Basic hospital (1.3%). Ages ranged from 16 to 19, with relatively equal representation. More than half were from urban municipalities (51.9%). Out of total; 16.9% were married who were all female participants (Table 3).

**Table 3:** Background characteristic of respondents.

| Variables | Background of participants | Number | Percentage (%) |
| --- | --- | --- | --- |
| Name of district | Surkhet | 18 | 11.7 |
|  | Banke | 20 | 13.0 |
|  | Pyuthan | 34 | 22.1 |
|  | Nuwakot | 17 | 11.0 |
|  | Parsa | 31 | 20.1 |
|  | Siraha | 34 | 22.1 |
| Type of Local Level | Municipality | 80 | 51.9 |
|  | Rural Municipality | 74 | 48.1 |
| Types of Health Facility | Basic Hospital | 2 | 1.3 |
|  | Primary Health Care Center | 10 | 6.5 |
|  | Health Post | 108 | 70.1 |
|  | Basic Health Care Center | 34 | 22.1 |
| Age of respondents | 16 year | 44 | 28.7 |
|  | 17 year | 40 | 26 |
|  | 18 year | 39 | 25.3 |
|  | 19 year | 31 | 20.1 |
| Education level of respondents | Basic education (Grade 1-8) | 55 | 35.7 |
|  | Secondary Education Examination | 62 | 40.3 |
|  | Higher Secondary (Grade 11-12) | 37 | 24 |
| Gender of respondent | Male | 47 | 30.5 |
|  | Female | 107 | 69.5 |
| Caste/ethnicity of respondents | Hill Brahmin | 7 | 4.5 |
|  | Hill Chhetri | 10 | 6.5 |
|  | Terai Brahmin and Chhetri | 5 | 3.2 |
|  | Terai other caste | 21 | 13.6 |
|  | Hill Dalit | 26 | 16.9 |
|  | Terai Dalit | 13 | 8.4 |
|  | Newar | 7 | 4.5 |
|  | Hill Janajati | 28 | 18.2 |
|  | Terai Janajati | 26 | 16.9 |
|  | Muslim | 9 | 5.8 |
|  | Others | 2 | 1.3 |
| Marital status of respondents | Married | 26 | 16.9 |
|  | Unmarried | 128 | 83.1 |
| Total |  | 154 | 100 |

The findings show that the respondents were generally educated, with representation across basic (35.7%), secondary (40.3%), and higher secondary (26%) education levels. One-third (30.5%) of sample were male adolescents. The background distribution also indicates substantial diversity in caste and ethnicity, with relatively higher representation from Janajatis (Hill: 18% and Terai: 17%) and Dalit (Hill: 17% and Terai: 8%) communities.

For the logistic regression analysis, several categories were merged to ensure adequate cell sizes and meaningful comparisons (Table 4). Basic hospitals and PHCC were combined into a single group. Hill Brahmin/Chhetri and Terai Brahmin/Chhetri were merged under “Brahmin/Chhetri.” Similarly, Hill Dalit and Terai Dalit were grouped as “Dalit,” and Newar, Hill Janajati, and Terai Janajati were merged into a single “Janajati” category. Muslim, Terai other castes, and other minor groups were combined under “Others.”

**Table 4:** Bivariate and multivariate logistic regression analysis of factors influencing health facility satisfaction on the day of visit.

| Variables |  | Bivariate Logistic Regression |  |  | Multivariate Logistic Regression |  |
| --- | --- | --- | --- | --- | --- | --- |
|  |  | Frequency | % | COR/ 95% CI (L-U)* | P- value | AOR/ CI |
| **Name of district** | Surkhet (ref) | 18 | 83.3 |  | .269 |  |
|  | Banke | 20 | 90 | 1.8 (0.27 – 12.23) | .550 | 2.10 (0.18-23.99) |
|  | Pyuthan | 34 | 100 | - | .998 | - |
|  | Nuwakot | 17 | 82.4 | 0.93 (0.16 – 5.42) | .883 | 1.23 (0.88-16.85) |
|  | Parsa | 31 | 83.9 | 1.04 (0.22 – 4.98) | .279 | 3.35 (376-29.78) |
|  | Siraha | 34 | 58.8 | 0.29 (0.07 – 1.18) | .521 | .51 (.063-4.05) |
| **Type of Local Level** | Municipality (ref) | 80 | 77.5 |  |  |  |
|  | Rural Municipality | 74 | 87.8 | 2.1 (0.88 – 5.02) | 0.47- | 1.9 (0. 457-7.91) |
| **Type of health facility** | PHCC and basic hospital (ref) | 12 | 66.7 |  | .384 |  |
|  | Health post | 108 | 82.4 | 2.34 (0.64 – 8.58) | .221 | 2.68 (.554-12.97) |
|  | Basic health care center | 34 | 88.2 | 0.77 – 18.39 | .704 | 1.52(.176-13.13) |
| **Age of respondents** | 16 years (ref) | 44 | 79.5 |  | .920 |  |
|  | 17 Years | 40 | 85 | 1.46 (0.47 – 4.54) | .562 | 1.50(.383-5.85) |
|  | 18 Years | 39 | 79.5 | 0.1 (0.34 – 2.90) | .954 | 1.04 (.255-4.25) |
|  | 19 Years | 31 | 87.1 | 1.74 (0.48 – 6.25) | .706 | 1.42 (0.23-8.84) |
| **Education level of respondents** | Basic education (Grade 1-8) (ref) | 55 | 81.8 |  | .235 |  |
|  | Secondary Education Examination | 62 | 85.5 | 1.31 (0.49 – 3.50) | .094 | 2.901 |
|  | Higher Secondary (Grade 11-12) | 37 | 78.4 | 0.81 (0.29 – 2.28) | .567 | 1.57 (.834-7.33) |
| **Gender** | Male (ref) | 47 | 83 |  |  |  |
|  | Female | 107 | 82.2 | 0.95 (0.38 – 2.36) | .511 | .68 (.221-2.11) |
| **Ethnicity** | Brahmin/Chhetri (ref) | 22 | 95.5 |  | .601 |  |
|  | Dalit | 39 | 87.2 | 0.32 (0.04 – 2.97) | .422 | .32 (.020-5.14) |
|  | Janajati | 61 | 77 | 0.16 (0.02 – 1.30) | .278 | .21 (.013-3.47) |
|  | Other ethnicity | 32 | 78.1 | 0.17 (0.02 – 1.50) | .204 | .14 (0.01-2.85) |
| **Marital status of respondents** | Married (ref) | 26 | 92.3 |  |  |  |
|  | Unmarried | 128 | 80.5 | 0.34 (0.08 – 1.55) | .429 | .45 (0.06-3.38) |
| **Total** |  |  | **82.5** |  |  |  |

\*L= Lower and \*U= Upper

Overall, 82.5% of adolescents reported being satisfied with the health services received on the day of their visit to health facility. Bivariate logistic regression analysis showed that none of the socio-demographic variables including age, gender, education, caste/ethnicity, marital status, place of residence, type of local government, or type of health facility were statistically significant predictors of adolescent satisfaction with the services.

However, several intermediate health system factors showed significant associations in the bivariate analysis. Adolescents who perceived the services as non-discriminatory had significantly higher odds of reporting satisfaction compared to those who did not (COR: 4.63, 95% CI: 1.94–11.05, p < 0.001). Similarly, those who reported that their confidentiality was maintained were more likely to be satisfied (COR: 5.39, 95% CI: 2.24–12.96, p < 0.001). The strongest crude association was observed for ease of obtaining an appointment with a health worker; adolescents who reported that it was easy to get an appointment had substantially higher odds of satisfaction (COR: 8.81, 95% CI: 3.53–32.00, p < 0.001). In contrast, availability of IEC materials and perceived satisfaction with adolescent participation were not significantly associated with satisfaction.

In the multivariate logistic regression model (Table 5), after adjusting for potential confounders, only two variables remained statistically significant. Adolescents who reported that the facility will keep their information confidential had higher odds of being satisfied with the services (AOR: 3.50, 95% CI: 1.19–10.37, p = 0.02). Likewise, ease of obtaining an appointment with a health worker remained a strong predictor of satisfaction (AOR: 6.28, 95% CI: 2.18–18.08, p < 0.001).

**Table 5:** Bivariate and multivariate logistic regression analysis of intermediate factors influencing health facility satisfaction.

| Variables |  | Client satisfaction with services received on the day of visit |  |  |  |  |
| --- | --- | --- | --- | --- | --- | --- |
|  |  | Satisfied % | Bivariate | Logistic | Multivariate | Logistic |
|  |  |  | Regression |  | Regression |  |
|  |  |  | p-value | COR, 95% CI(L-U) | p-value | AOR, 95% CI(L-U) |
| Availability of IEC materials for reading and takeaway | No/Don't know (ref) | 78.9 |  |  |  |  |
|  | Yes | 91.1 | 0.079 | 2.74 (0.89-8.44) | 0.87 | 1.12 (0.28-4.53) |
| Health facility provides non-discrimination services | Disagree (ref) | 64.3 |  |  |  |  |
|  | Agree | 89.3 | <0.001*** | 4.63 (1.94-11.05) | 0.85 | 1.13 (0.34-3.79) |
| Health facility maintaining confidentiality of information | No (ref) | 62.8 |  |  |  |  |
|  | Yes | 90.1 | <0.001*** | 5.39 (2.24-12.96) | 0.02* | 3.50 (1.19-10.37) |
| Level of satisfaction towards adolescent participation | Dissatisfaction (ref) | 80.5 |  |  |  |  |
|  | Satisfied | 95.2 | 0.131 | 4.86 (0.62-37.89) | 0.83 | 1.28 (0.13-12.59) |
|  | Difficult (ref) | 52.9 |  |  |  |  |
| **Taking**<br>**appointment**<br>**with health**<br>**workers** | Easy | 90.8 | <0.001*** | 8.81<br>(3.53-32.00) | <0.001*** | 6.28 (2.18-18.08) |

Other variables, including perceived non-discrimination, availability of IEC materials, and adolescent participation, were not statistically significant in the adjusted model. This suggests that their initial associations observed in the bivariate analysis may be explained by confounding or overlapping effects with other variables included in the model.

### Qualitative findings

The FGDs provided detailed insights into adolescents’ perceptions of health services, particularly regarding confidentiality, provider behavior, interactions and appointment processes, and their participation in health-system decision-making. The analysis focused on key themes aligned with the study framework, including accessibility, acceptability, provider behavior, communication, and adolescent participation in health system processes. The findings are presented thematically to illustrate adolescents lived experiences and to enrich interpretation of the quantitative results.

### Theme 1: Accessibility of services

In the FGDs, adolescents reported that frequent stock outs of medicines undermined their trust in public health facilities, pushing them to seek care from private providers, who were perceived as more reliable but required additional financial costs and travel. Stock outs of condoms and the absence of condom boxes further limited their ability to access contraception easily in the public health facilities.

“*We go to private clinic when we need condoms and other medicine where we can get them immediately as per our needs and preferences, without being asked personal questions. There is no need to disclose anything about us, and we can return quickly from there. There is no worry about the stock out of the medicine*”. - *male adolescent group, FGD in Surkhet*.

Many adolescent boys reported preferring private clinics over public health facilities, as they could obtain condoms with minimal interaction with service providers, thereby reducing discomfort and the risk of embarrassment.

“*We just go private clinic for the condom, disappear quickly from where there is no fear of being identified” male adolescent group, FGD in Siraha*

*“We want our public health facility be like, where we can grab the thing without talking much to them and come back”. – male adolescent group, FGD in Surkhet*.

Girls in Madhesh Province (Siraha district) shared that unmarried adolescents particularly seeking contraceptive services are often asked by health workers to bring a parent or family member for consent. This practice created a major barrier for them, as it reflected prevailing sociocultural norms that view contraceptive use and, by implication, sexual activity as unacceptable for unmarried girls. As a result, many of them felt discouraged from seeking the services they needed from nearest service facility.

*“Health workers ask us to come with parents or family member to take the any kind of health service including contraceptive services” – unmarried female adolescent group, FGD in Siraha*.

An adolescent girls’ group in Parsa reported experiences similar to those in Siraha, describing judgmental attitudes from health service providers that made unmarried to feel uncomfortable and discouraged from seeking contraceptive services.

“*It is not difficult for married girls to take contraceptive service from health post, but for unmarried girls it is very challenging. Health workers ask several questions and perceive in different ways. With the fear of being judged we just go the private clinic”. - female adolescent group, FGD in Parsa*.

Adolescents also reported challenges in accessing services when the available health workers were of the opposite gender. Across districts, boys explained that they are unable to discuss reproductive health concerns with female providers, which often resulted in delaying or avoiding services. This gender-related discomfort similarly affected girls, who expressed hesitation when only male health workers were present in the facility.

“*I had a problem with my private organ but did not go to health post for a check-up. It later resolved on its own without any treatment. I did not seek care because only female health workers were available.”* — *male adolescent group, FGD in Pyuthan*.

“*If there were only male health workers, I would rather ask my mother to get the medicine from there*”. — *female adolescent group, FGD in Pyuthan*.

### Theme 2: Acceptability of services

Confidentiality emerged as a major barrier to adolescents’ use of contraceptive services in the FGDs, closely aligning with the conceptual framework that identifies confidentiality as a key determinant of service satisfaction. Adolescents reported discomfort seeking sensitive services particularly contraception, from the facilities located within their own communities, where both health workers and visitors were often personally known to them. They feared that information about their visits would be disclosed to family members or others in the community, potentially exposing them to gossip, blame, or social sanctions.

*“If we request condoms at our nearby public health facility, the information often reaches to our family members before we arrive at home”* – *male adolescents’ group, FGD Banke*.

Adolescents noted that the physical layout of the public health facilities compromised privacy, particularly due to open and highly visible waiting areas where they felt vulnerability of being exposed to scrutiny from other clients, including relatives. Although waiting times were not necessarily long, the fear of being seen deterred many adolescents from seeking services.

“*Private discussions with health workers are only possible when the providers are free, often requiring long waits*. S*ometimes it is possible either at late afternoon or before start of the clinic*”. — *female adolescent group, FGD in Siraha*.

These concerns highlight confidentiality as a critical intermediate factor shaping adolescents’ satisfaction and their willingness to use contraceptive services in public health facilities. They illustrate how perceived risks to privacy generate fear, discourage service uptake, and ultimately influence overall satisfaction with health services. In Siraha and Parsa (Madhesh province), adolescents reported using coded language such *“Suraksha”* (safety in Nepali language) to request condoms, as openly mentioning condoms in public remain socially taboos within their communities.

“*If the condom box is empty at health facility, we usually leave and try again another time. We don’t feel comfortable asking them for condoms in person. If we ask, they label us as boy of inappropriate behaviors”.* — *male adolescent group, FGD in Parsa*.

### Theme 3: Friendly and Non-Judgmental

Health workers’ behaviour emerged as another important health system factor influencing adolescents’ trust, comfort, and satisfaction with health services. Adolescents described experiences of judgmental attitudes, intrusive or unnecessary questioning, and a lack of empathy, particularly in relation to SRH services. Many perceived the questions asked during consultations as embarrassing, disrespectful, and discouraging, especially among unmarried adolescents seeking SRH care.

*“Some health workers look at us with bad eye. If we encounter such behavior, we never back to that health facility for services to avoid being judged and treated differently at health facility”.* — *female adolescent group, FGD in Siraha*

*“The health workers chased us away from health facility if we ask for condom saying they would tell our parents”* — *male adolescents group, FGD in Banke*.

### Theme 4: Communication

Adolescents described several communications-related challenges in their interactions with health workers when seeking services. During visits for general outpatient services, they reported that consultations were brief, clinical examinations were superficial, and the medicines provided were limited to a narrow set of basic drugs, most commonly paracetamol or iron tablets. They were often asked highly personal and sensitive questions. Participants perceived these inquiries as intrusive, unnecessary, and embarrassing, particularly when asked in the presence of other clients. These communication practices caused discomfort, discouraged open discussion of reproductive health issues, and reduced adolescents’ willingness to seek contraceptive services at public health facilities.

*“They even asked about the name of our parent, in such case we have to disappear from there instead of taking the services”* — *male adolescent group, FGD in Surkhet*.

Some adolescents felt they were not prioritized for services, and communication barriers further complicated interactions, particularly in the Terai where many adolescents primarily speaking local languages and struggled with Nepali speaking service providers.

*“Sometimes health workers do not speak the local language as they are from other community, and rural girls like us cannot communicate in Nepali language clearly to receive the service. We have to come with visitors for communication, and in that situation, it is not possible to share our problems clearly with health workers because it breaches our privacy in front of our family or relatives”* — *female adolescent group, FGD in Siraha*.

Regarding the use of IEC materials, participants reported limited availability of such resources at health facilities. In all districts, adolescents noted that there were insufficient materials to read while waiting for services or to take home for later reference. As a result, they relied on alternative sources, including books, the internet, and social media platforms such as Facebook, TikTok, ChatGPT, and YouTube to obtain information about their reproductive health.

“*I find it more convenient to seek information through ChatGPT rather than consulting IEC materials or books to understand my reproductive health, as it is quicker and easier*.” — *female adolescent group, FGD in Pyuthan*.

### Theme 5: Participation and engagement

Adolescents reported minimal participation in local health system governance structures, including planning and decision-making processes. Most were unaware that an adolescent representative is formally included in the Health Facility Operation and Management Committee (HFOMC). Among the respondents, none had ever been invited to HFOMC meetings or consultations related to planning, budgeting, or service improvements to adolescent health programs. Participants from Siraha, Banke, and Nuwakot further explained that committee chairpersons often invite adolescents who were close relatives of HFOMC members, thereby limiting broader and more equitable adolescent engagement in health system governance.

“*Health facility only calls the elders or our parents in the meeting. They don’t think adolescent like us need to be invited at the meeting of health facility*” — *male adolescent group, FGD in Parsa*

Information about annual community-level planning and budget meetings was perceived as poorly disseminated, and participants raised concerns about limited transparency and accountability within health facilities. They also noted that community and health issues were often politicized, which constrained opportunities for meaningful youth participation. This lack of engagement contributed to a sense of detachment among adolescents and reduced their motivation to seek services.

### Integration of quantitative and qualitative data on client satisfaction

The integration of quantitative and qualitative findings (Table 6) offered a richer understanding of adolescents’ satisfaction with public health services[20]. While the quantitative data indicated generally high levels of overall satisfaction, particularly with general outpatient services, the qualitative insights highlighted a more nuanced picture regarding contraceptive services. Many adolescents expressed significant dissatisfaction, especially concerning issues of privacy, provider attitudes and behaviors, and the challenges associated with seeking SRH services and information. This suggests that the high satisfaction scores reported during exit interviews might have been influenced by social desirability bias.

**Table 6:** Integration quantitative and qualitative findings.

| **Quantitative score/**<br>**finding** | **Qualitative finding** | **Meta-inference** |
| --- | --- | --- |
| 82.5% participants satisfied with the services. | The adolescent expressed dissatisfaction with health services mentioning that there is issues privacy, confidentiality, discriminatory service, judgmental | Inconsistence finding because of possible social desirability bias. |
|  | attitude of health worker is judgmental (**Theme 1-5**). |  |
| Statistically no significant association between IEC materials and satisfaction with health facility (AOR: 1.12 (0.28-4.53)) | **Theme 4:** The adolescent felt that there are no enough IEC materials to read and take-home at health facilities. Moreover, they rely on alternative sources for the information and feel more convenient. | The findings are consistence. IEC materials do not have influence to satisfaction with health facility. |
| Statistically no significant association between non-discriminatory service and satisfaction with health facility (AOR:1.13). | **Theme 1:** Felt discriminated in access to service based on marital status and age. | Quantitative finding did not detect a statistical relationship, but qualitative finding reveals subsisted discriminatory experiences. |
| Statistically significant association between confidentiality of information and health facility (AOR: 3.50). | **Theme 2:** The participants felt that is high possibility of breezing privacy if they seek contraceptive service from public health facilities.<br>**Theme 3:** Health workers are judgmental in providing SRH service.<br>**Theme 4:** Privacy and confidentiality of information are not acknowledged by health workers. | Both findings indicate that confidentiality is a determinant of health service use and satisfaction with health facility. |
| No statistically significant between adolescent participation and health facility (AOR: 1.28). | **Theme 5:** Participants of adolescents in health system is not adequately recognized and ensured at local level. | Weak or absent participation of adolescent in health system may explain why no measurable effect is identified. |
| Significant association between taking appointment with health | **Theme 1:** not willing to take appointment with health worker of public health facility with fear of breezing the confidentiality. | Result is consistence, ease of appointment with the health is the determinants of |
| workers and health facility<br>(AOR: 6.28). | **Theme 2:** need to wait long to talk to<br>health worker in private. | satisfaction with health<br>facility. |

Key determinants identified in the quantitative analysis, such as the confidentiality of information and ease of access to healthcare providers, were strongly supported by qualitative evidence. Adolescents consistently highlighted the importance of privacy, trust, and approachable communication in shaping their experiences with services and service seeking behaviour. The gap between overall service satisfaction and specific concerns regarding contraceptive services underscores the advantages of employing a mixed-methods approach. This methodology reveals context-specific barriers that adolescents may encounter, which may not be fully captured through quantitative measures alone.

## Discussion

While the quantitative findings indicate a high level of reported satisfaction (82.5%), the integrated analysis demonstrates that this apparent satisfaction hides important gaps in adolescents lived experiences, particularly in relation to SRH services. The divergence between quantitative and qualitative findings highlights the limitations of relying solely on quantitative satisfaction measurement, especially in relation to sensitive services such as contraception, where social desirability bias and contextual constraints may influence responses.

This study demonstrates that adolescents’ satisfaction is less shaped by socio-demographic characteristics more by the health system factors in line with finding of previous conducted in Nepal[30]. This finding is also consistent with evidence from countries such as Ghana and Tanzania, where structural and interpersonal dimensions of care, particularly privacy, respectful communication, and timely access have been shown to be more influential than individual characteristics in shaping adolescents’ healthcare experiences [31]. Interpreted through the Andersen Behavioral Model, these findings emphasize the dominant role of intermediators in determining both satisfaction and subsequent service utilization.

In the adjusted analysis, ease of accessing providers and assurance of confidentiality emerged as the strongest predictors of satisfaction. Adolescents expressed a clear preference for quick, discreet, and low-interaction services, reflecting a desire to minimize exposure and social risk. Previous studies in Nepal have similarly shown that easily accessible providers enhance adolescents’ ability to seek services and obtain contraceptives when needed, making services more responsive to their needs [17,18,30]. This relationship is reciprocal: when services are available at convenient times, access improves, and when providers are easily accessible, adolescents are more likely to use services when the need arises. Consistent with these findings, the availability of SRH services, including contraception, at convenient times has also been identified as a positive factor influencing service utilization in other developing countries, such as Ethiopia and Kenya. [32,33].

The central role of confidentiality identified in this study aligns strongly with global evidence positioning privacy as a cornerstone of adolescent-responsive health systems [34,35]. Moreover, this is also aligned with finding from earlier study conducted in Nepal [17]. When confidentiality is assured, adolescents are more likely to seek care, disclose their actual problems, and using services. Conversely, even perceived risks to confidentiality can act as powerful deterrents.

The qualitative findings provide critical explanatory depth to these associations. Adolescents describe public health facilities as socially exposed spaces where privacy is difficult to maintain, particularly due to open waiting areas, familiarity with providers, and the risk of disclosure of information to family or community members. These concerns were especially pronounced for contraceptive services, which are closely linked to social norms surrounding sexuality. Similar evidence from Ethiopia, Ghana and Tanzania shows that perceived or actual breaches of confidentiality significantly reduce adolescents’ willingness to seek SRH services, often pushing them toward private or informal providers where anonymity is better preserved [33,36,37].

In addition to confidentiality, provider behavior emerged as a critical determinant of adolescents’ experiences. Reports of judgmental attitudes, intrusive questioning, and perceived discrimination particularly among unmarried girls highlight how social and gender norms are reproduced within health service delivery. Evidence from different developing countries from Sub Sharan region and India similarly demonstrates that provider bias and moralistic attitudes toward adolescent sexuality discourage service utilization and undermine trust in public health systems[38–41]. In the context of Nepal, these barriers are further compounded by cultural norms that stigmatize premarital sexual activity, making it particularly difficult for unmarried adolescents to access contraceptive services[42,43].

An important structural factor identified in this study relates to the legal and social context of marriage. The legal age of marriage in Nepal may unintentionally create systemic barriers, though provision of SRH information and service is available irrespective of marital status. Some married adolescents may misreport their age to avoid legal implications. This not only affects accurate identification and inclusion of adolescents in services but also reflects broader tensions between legal frameworks and social realities. Similar discrepancies between legal norms and adolescent behaviors have been observed in India [40], where early marriage persists despite legal restrictions, influencing access to and reporting of reproductive health services.

The finding that availability of IEC materials was not associated with satisfaction suggests that the presence of information alone is insufficient to influence adolescents’ service experiences. This aligns with emerging global evidence indicating that adolescents increasingly rely on digital platforms and social medias and peer networks for SRH information [44,45]. This reflects a broader paradigm shift from traditional, facility-based information dissemination toward more accessible, user-driven digital sources. For many adolescents, seeking information through health workers may feel less convenient or less private, making it a less preferred option. Therefore, it is essential for health systems to recognize these evolving preferences and adapt accordingly. Aligning to our finding, studies from India have also shown that while facility-based IEC materials remain important [46], their effectiveness depends on relevance, accessibility, and privacy of engagement factors that may currently be inadequate in many public health settings.

Meaningful participation of adolescent is essential for delivering responsive, developmentally appropriate care that places adolescents at the center of service design and delivery[47]. However, the study also highlights the limited participation of adolescents in health system governance and decision-making processes. Despite policy provisions for youth engagement, adolescents reported minimal awareness of or involvement in mechanisms such as the HFOMC. This disconnect between policy intent and practice is consistent with findings from Nepal and other LMICs, where community participation structures often remain tokenistic and dominated by local elites. Lack of meaningful adolescent engagement reduces the responsiveness of services and perpetuates gaps between service design and user needs.

Overall, this study demonstrates that adolescents’ satisfaction and utilization of health services are shaped by a complex interplay of health system factors and socio-cultural norms. Public health facilities, as currently structured, may not adequately meet the unique needs of adolescents, particularly for SRH services. The findings strongly suggest that improving adolescent health outcomes requires moving beyond a service availability approach toward a more adolescent-centered model of care, one that prioritizes confidentiality, respectful and non-judgmental interactions, accessible service delivery, and meaningful adolescent engagement.

## Conclusion

This study demonstrates that reported satisfaction with public health services does not fully capture adolescents’ actual experiences, particularly in relation to SRH services. Despite seemingly high satisfaction, adolescents are not the primary users of public health facilities, indicating important gaps in service accessibility and acceptability. The findings highlight that health system factors, especially confidentiality and ease of access to providers play a central role in shaping adolescents’ experiences and their willingness to seek care. At the same time, socio-cultural norms, stigma, and judgmental provider attitudes continue to act as a barrier, particularly for unmarried adolescents seeking contraceptive services. These findings underscore the need to reorient public health services toward a more adolescent-centered approach that prioritizes privacy, respectful care, and accessible service delivery. Strengthening these aspects is essential to improve service utilization and advance adolescent sexual and reproductive health outcomes in Nepal.

## Limitation

This study has several limitations. Although, the study employed a sequential explanatory design; however, the quantitative data were collected at a single point in time, which may limit the ability to capture temporal variations. Client exit interviews were conducted only among adolescents who visited selected health facilities on days when enumerators were present. In addition, the study was limited to public health facilities, with private health facilities not included, which may constrain the generalizability of the findings across different service delivery settings.

Despite these limitations, the study was strengthened by methodological triangulation, as quantitative findings were complemented and validated through qualitative data, enhancing the comprehensiveness and interpretability of the results. The selection of districts with a high burden of teenage pregnancy further contributed to the relevance and contextual strength of the study.

## Data Availability

The data underlying the findings of this study contain sensitive participant information and cannot be made publicly available due to ethical restrictions. Data are available from the corresponding author upon reasonable request and with approval from the Nepal Health Research Council.

## Abbreviation

AFHS: Adolescent friendly health service
ASRH: Adolescent Sexual and Reproductive Health
FGD: Focus groups discussion
HFOMC: Health facility operation and management committee
IEC: Information education and communication
PHCC: Primary health care center
SRH: Sexual reproductive health
SRHR: Sexual and reproductive health and rights
SPSS: Statistical package for the social sciences

## Acknowledgements

The authors would like to acknowledge the support of the Acting Dean of the School of Arts, Kathmandu University, Associate Professor Dr. Uddab Pyakurel; Associate Professor Dr. Chandra Lal Pandey; Associate Professor Dr. Binayak Krishna Thapa, Head of the Department of Development Studies, School of Arts, Kathmandu University; Prof. Dr. Ramesh Adhikari, Tribhuvan University; Sujan Karki and Mr. Abhisek Karna for their valuable review support.

## Authors Contributions

**Conceptualization:** Santa Kumar Dangol.

**Data collection:** Santa Kumar Dangol, Adweeti Nepal.

**Data curation:** Santa Kumar Dangol, Adweeti Nepal.

**Formal analysis:** Santa Kumar Dangol, Adweeti Nepal.

**Software:** Santa Kumar Dangol.

**Methodology (coding and transcription):** Santa Kumar Dangol, Adweeti Nepal.

**Writing – original draft:** Santa Kumar Dangol, Adweeti Nepal.

**Writing – review & editing:** Santa Kumar Dangol, Adweeti Nepal, Megh Raj Dangal, Ramesh Adhikari, Sujan Babu Marahatta..

**Supervision and validation:** Megh Raj Dangal.

## Declaration on use of Artificial Intelligence tool

The authors acknowledge the use of ChatGPT for language refinement and improvement of writing clarity. In addition, Grammarly was used for grammar checking and editing. Grammarly was accessed through the LMU student account of Adweeti Nepal, while ChatGPT was used via its online platform without institutional account access. Both tools were used solely to support language quality, and all intellectual content, analysis, and interpretations remain entirely the responsibility of the authors.

## Footnotes

1 Plain and flat geographical region in southern part of Nepal

## References

1. WHO. Global Accelerated Action for the Health of Adolescents (AA-HA!): Guidance to Support Country Implementation. World Health Organization; 2023.

2. Adolescents Statistics - UNICEF DATA. [cited 9 Mar 2026]. Available: https://data.unicef.org/topic/adolescents/overview/

3. UNICEF. Early childbearing and teenage pregnancy rates by country - UNICEF DATA. Nov 2024 [cited 31 Jul 2025]. Available: https://data.unicef.org/topic/child-health/early-childbearing/

4. WHO. WHO Guidelines on Preventing Early Pregnancy and Poor Reproductive Outcomes. World Health Organization. 2011; 192. Available: http://whqlibdoc.who.int/publications/2011/9789241502214_eng.pdf?ua=1

5. Wells JC, Noviyanti Q, Marphatia AA, Rougeaux E. Early Marriage, Preterm Birth, and School Dropout: An Intergenerational Cycle of Risk? American Journal of Human Biology. 2025;37. doi:10.1002/ajhb.70177

6. Sekine K, Hodgkin ME. Effect of child marriage on girls’ school dropout in Nepal: Analysis of data from the Multiple Indicator Cluster Survey 2014. PLoS One. 2017;12. doi:10.1371/journal.pone.0180176

7. National Statistics Office. National Population and Housing Census 2021 Adolescents and Youth in Nepal. 2025. Available: www.nsonepal.gov.np

8. Ministry of Health and Population; New ERA; and ICF. Nepal Demographic and Health Survey 2022. Kathmandu, Nepal: Ministry of Health and Population (Nepal). Kathmandu, Nepal: Ministry of Health and Population (Nepal); 2022.

9. Ministry of Health and Population, Nepal Statistic Office. National Population and Housing Census 2021: Nepal Maternal Mortality Study 2021. Kathmandu: Ministry of Health and Population; National Statistics Office; 2022.

10. Subedi R, Jahan I, Baatsen P. Factors Influencing Modern Contraceptive Use among Adolescents in Nepal. J Nepal Health Res Counc. 2018;16: 251–256. doi:10.3126/jnhrc.v16i3.21419

11. Dagnew T, Tessema F, Hiko D. Original Article Health and Reported Satisfaction Among Adolescents in Dejen District, Ethiopia: A Cross-Sectional Study. Ethiop J Health Sci. 2015;25: 17–28.

12. Sharma M, Khatri B, Amatya A, Subedi N, Upadhyaya DP, Sapkota BP, et al. Utilization of adolescent friendly health services and its associated factors among higher secondary students in mid-western Himalayan mountainous district of Nepal. PLOS Global Public Health. 2023;3. doi: 10.1371/journal.pgph.0001616

13. D’Souza P, Bailey J V., Stephenson J, Oliver S. Factors influencing contraception choice and use globally: a synthesis of systematic reviews. European Journal of Contraception and Reproductive Health Care. 2022;27: 364–372. doi:10.1080/13625187.2022.2096215

14. Family Welfare Division. National Adolescent Health and Development Strategy 2018. Kathmandu, Nepal: Government of Nepal, Ministry of Health and Population, Department of Health Services, Family Welfare Division; 2018.

15. Family Welfare Division. Adolescent Friendly Health Service Operation Guideline 2022. Kathmandu, Nepal: Government of Nepal, Ministry of Health and Population, Department of Health Service, Family Welfare Division, Nepal; 2022.

16. Government of Nepal. The Right to Safe Motherhood and Reproductive Health Act, 2075 (2018). Nepal Law Commission; 2018 pp. 1–13.

17. Pandey PL, Seale H, Razee H. Exploring the factors impacting on access and acceptance of sexual and reproductive health services provided by adolescent-friendly health services in Nepal. PLoS One. 2019;14. doi:10.1371/journal.pone.0220855

18. Napit K, Shrestha KB, Magar SA, Paudel R, Thapa B, Dhakal BR, et al. Factors associated with utilization of adolescent-friendly services in Bhaktapur district, Nepal. J Health Popul Nutr. 2020;39: 2. doi:10.1186/s41043-020-0212-2

19. USAID, PACE, PRB. Adolescent Fertility Rate (AFR) in Nepal: Better evidence to drive more impactful allocation of family planning resources. 2024. Available: http://adolescentfertility.dds4dev.org/

20. John W. Creswell, J. David Creswell. Research Design: Qualitative, Quantitative and Mixed Methods Approaches. Sixth. SAGE Publications, Inc.; 2023.

21. Andersen RM. Revisiting the Behavioral Model and Access to Medical Care: Does it Matter? 1995;36: 1–10.

22. Cochran WG. Sampling techniques. Third Edition. Harvard University: Wiley; 1977.

23. Nasser RN. Using the Spreadsheet to Understand Random Sampling Procedures in Relation to the Central Limit Theorem. J Math Stat. 2008;4: 168–173.

24. Israel GD. Determining Sample Size 1. Available: http://edis.ifas.ufl.edu.

25. Naeem M, Ozuem W, Howell K, Ranfagni S. A Step-by-Step Process of Thematic Analysis to Develop a Conceptual Model in Qualitative Research. Int J Qual Methods. 2023;22: 1–18. doi:10.1177/16094069231205789

26. Braun V, Clarke V. Using thematic analysis in psychology. Qual Res Psychol. 2006;3: 77–101. doi:10.1191/1478088706qp063oa

27. Gautam L, Maharjan A, Bhattarai H, Gautam S. Availability and utilization of sexual and reproductive health services among adolescents of Godawari Municipality, Nepal: A cross-sectional study. PLOS Global Public Health. 2025;5. doi:10.1371/journal.pgph.0003282

28. Birhanu Z, Tushune K, Jebena MG. Sexual and Reproductive Health Services Use, Perceptions, and Barriers among Young People in Southwest Oromia, Ethiopia. Ethiop J Health Sci. 2018;28: 37–48. doi:10.4314/EJHS.V28I1.6

29. Tetui M, Ssekamatte T, Akilimali P, Sirike J, Fonseca-Rodríguez O, Atuyambe L, et al. Geospatial Distribution of Family Planning Services in Kira Municipality, Wakiso District, Uganda. Front Glob Womens Health. 2020;1: 1–10. doi:10.3389/fgwh.2020.599774

30. Pahari S, Acharya SR, Pokhrel A, Upadhyay JP, Parajuli S, Pudasainee M, et al. Adolescent-friendly health services in Nepal: usage and key determinants. BMC Health Serv Res. 2025;25: 1000. doi:10.1186/s12913-025-13157-y

31. Hutchinson PL, Do M, Agha S. Measuring client satisfaction and the quality of family planning services: A comparative analysis of public and private health facilities in Tanzania, Kenya and Ghana. BMC Health Serv Res. 2011;11. doi:10.1186/1472-6963-11-203

32. Kinaro J, Kimani M, Ikamari L, Ayiemba EHO. Perceptions and Barriers to Contraceptive Use among Adolescents Aged 15 - 19 Years in Kenya: A Case Study of Nairobi. Health N Hav. 2015;07: 85–97. doi:10.4236/health.2015.71010

33. Habtu Y, Kaba M, Mekonnen H. What do service providers in Southern Ethiopia say about barriers to using youth-friendly sexual and reproductive health services for adolescents?: Qualitative study. Reprod Health. 2021;18. doi:10.1186/s12978-021-01092-0

34. Ford CA, English A, Dowshen N, Rogers CG. Confidentiality in adolescent health care. Health Promotion for Children and Adolescents. 2016;135: 347–370. doi:10.1007/978-1-4899-7711-3_17

35. Pampati S, Liddon N, Dittus PJ, Adkins SH, Steiner RJ. Confidentiality Matters but How Do We Improve Implementation in Adolescent Sexual and Reproductive Health Care? Journal of Adolescent Health. 2019;65: 315–322. doi:10.1016/j.jadohealth.2019.03.021

36. Report of an adolescent health services barriers assessment (AHSBA) in the United Republic of Tanzania With a Focus on Disadvantaged Adolescents The United Republic of Tanzania Ministry of Health, Community Development, Gender, Elderly and Children.

37. Nartey EB, Babatunde S, Okonta KE, Kotoh AM, Amoadu M, Abraham SA, et al. Prevalence and barriers to the utilization of adolescent and youth-friendly health services in Ghana: systematic review and meta-analysis. Reproductive Health. BioMed Central Ltd; 2025. doi:10.1186/s12978-025-02010-4

38. Lateef MA, Pillay JD. Mapping research evidence on the use of contraceptives among adolescent girls in Africa: a scoping study. BMC Public Health. 2026. doi:10.1186/s12889-026-26251-5

39. Meek C, Mulenga DM, Edwards P, Inambwae S, Chelwa N, Mbizvo MT, et al. Health worker perceptions of stigma towards Zambian adolescent girls and young women: a qualitative study. BMC Health Serv Res. 2022;22. doi:10.1186/s12913-022-08636-5

40. Shukla A, Kumar A, Mozumdar A, Acharya R, Aruldas K, Saggurti N. Restrictions on contraceptive services for unmarried youth: a qualitative study of providers’ beliefs and attitudes in India. Sex Reprod Health Matters. 2022;30. doi:10.1080/26410397.2022.2141965

41. Mbadu Muanda F, Gahungu NP, Wood F, Bertrand JT. Attitudes toward sexual and reproductive health among adolescents and young people in urban and rural DR Congo. Reprod Health. 2018;15. doi:10.1186/s12978-018-0517-4

42. Wasti C, Simmons SP, Limbu R, Chipanta N, Haile S, Velcoff L. USide-Effects and Social Norms Influencing Family Planning Use in Nepal. Kathmandu Univ Med J. 2017.

43. Angdembe MR, Sigdel A, Paudel M, Adhikari N, Bajracharya KT, How TC. Modern contraceptive use among young women aged 15-24 years in selected municipalities of Western Nepal: Results from a cross-sectional survey in 2019. BMJ Open. 2022;12. doi:10.1136/bmjopen-2021-054369

44. Saha R, Paul P, Yaya S, Banke-Thomas A. Association between exposure to social media and knowledge of sexual and reproductive health among adolescent girls: evidence from the UDAYA survey in Bihar and Uttar Pradesh, India. Reprod Health. 2022;19. doi:10.1186/s12978-022-01487-7

45. Waldman L, Ahmed T, Scott N, Akter S, Standing H, Rasheed S. “We have the internet in our hands”: Bangladeshi college students’ use of ICTs for health information. Global Health. 2018;14. doi:10.1186/s12992-018-0349-6

46. Bahl D, Bassi S, Maity H, Krishnan S, Dringus S, Mason-Jones A, et al. Compliance of Adolescent Friendly Health Clinics with National and International Standards: Quantitative findings from the i-Saathiya study. BMJ Open. 2024;14. doi:10.1136/bmjopen-2023-078749

47. Wigle J, Paul S, Birn AE, Gladstone B, Braitstein P. Youth participation in sexual and reproductive health: policy, practice, and progress in Malawi. Int J Public Health. 2020;65: 379–389. doi:10.1007/s00038-020-01357-8

